# Type 2 diabetes in Norway 2009–2021: Have declining incidence rates continued?

**DOI:** 10.1101/2025.09.24.25336539

**Authors:** Lars Bakke Hindenes, Paz Lopez-Doriga Ruiz, German Tapia, Inger Johanne Bakken, Elisabeth Qvigstad, Hanne L. Gulseth, Lars C. Stene

## Abstract

**Aims/hypothesis:** To investigate updated trends in the incidence and prevalence of diagnosed type 2 diabetes in Norway by age, sex, country of birth and education.

**Methods:** A nationwide cohort study using registries on primary and specialist healthcare, dispensed drugs, and demographic factors in Norway. We analysed incidence trends 2009-2021 using Poisson regression and JoinPoint.

**Results:** During 2009–2021, 195 935 incident type 2 diabetes cases were identified. After a decline in incidence 2009–2014, the incidence was largely stable over time, with a suggestive upward trend 2019–2021. The overall incidence rates were 651 per 100 000 person-years in 2009 and 530 per 100 000 in 2021. There were 165 432 prevalent cases of type 2 diabetes in 2009 (5.5% of the population), consistently increasing throughout the study period to a peak at 259 017 (7.5%) in 2021. The time trends were largely consistent across age, sex, country of birth and education. Inhabitants with lower education and born in Asia or Africa had substantially higher incidence and prevalence than those with higher education and born in Norway or other continents, respectively.

**Conclusions/interpretation:** The previously described decline in incidence of diagnosed type 2 diabetes during 2009– 2014 was not sustained, and prevalence continued to increase throughout 2009–2021.

**Research in context:** *What is already known about this subject?:*

- The incidence rate of type 2 diabetes in Norway and several other countries decreased from 2009–2014, despite an ongoing increase in prevalence.

*What is the key question?:*

- Has the decrease in incidence rate of type 2 diabetes in Norway continued after 2014?

*What are the new findings?:*

- The incidence rate of type 2 diabetes in 30–89-year-old residents in Norway was relatively stable from 2015–2021, with a suggestive increase from 2019–2021, largely consistent by sex, age, education and country of birth. The prevalence continued to increase.

*How might this impact on clinical practice in the foreseeable future?:*

- Both incidence and prevalence are important for shorter term health care planning in an ageing population and to understand time trends.

## Introduction

While the prevalence of type 2 diabetes has increased globally over several decades, the incidence of type 2 diabetes appears to stabilise or decrease in many high-income countries, including Norway, after 2010 [1, 2]. We reported a declining incidence of type 2 diabetes and a concomitant increasing prevalence during 2009–2014 in Norway [3].Studies from other European countries showed similar declining incidence trends [4-9].

Type 2 diabetes is more common among individuals with higher body mass index, older age, and lower socioeconomic status [4, 5]. Given these associations, and that obesity prevalence has risen in most countries [6, 7], the observed decline in type 2 diabetes incidence was unexpected.

Prevalence of type 2 diabetes is influenced by incidence and disease duration. As disease duration may be increased by both earlier detection and extended survival, prevalence may increase because of more intensive case-finding or screening, as well as extended survival for instance due to better treatment [5]. Therefore, changes in prevalence will naturally lag changes in incidence, and will not necessarily show the same trends. Few previous studies have investigated type 2 diabetes incidence stratified by individual data on country of birth and socioeconomic status, although some have used area-based indices [8, 9]. To understand the developing time trends and disease burdens, we investigated the incidence and prevalence trends of type 2 diabetes in Norway by age, sex, country of birth and education, to assess whether the previously reported declining incidence [3] continued 2015–2021.

## Methods

### Study participants and data sources

We included all individuals registered as being alive, living in Norway, and aged 30–89 in at least one of the study years 2009–2021. Individual-level data from population based nationwide registries were linked: Statistics Norway (SSB, population and education), the Norwegian Prescribed Drug Registry (NorPD, dispensed drug ATC codes, 1 January 2004–1 June 2022), the Norwegian Patient Registry (NPR, use of secondary health care, ICD-10 codes, 1 January 2008–31 December 2022) and the Norwegian Control and Payment of Health Reimbursements Database (KUHR, use of primary health care ICPC-2 codes, 1 January 2006–1 June 2022). These data sources are described in more detail at https://helsedata.no/en/. We used available data from both before and after the study period: earlier data (“look-back period”) to exclude prevalent diabetes from being counted as incident, and data after 2021 (5 or 12 months in different registries) to allow for a second entry in a registry (see next paragraph). This study is part of the DIANOREG project (Population-based register study of diabetes in Norway). The study was approved by the Regional Ethical Committee (REK 28748 (2019/1175)).

### Outcome: Incidence and prevalence of diagnosed type 2 diabetes

The primary outcome was incidence rate of diagnosed type 2 diabetes, and the secondary outcome was prevalence of type 2 diabetes, in each of the study years 2009–2021. As in our previous publication [3], type 2 diabetes was defined as having either (1) at least one registration of a type 2 diabetes diagnosis (ICPC-2 code T90 or ICD-10 code E11) combined with at least one dispensed non-insulin glucose-lowering drug (ATC codes starting with A10B), or (2) at least two separate registrations of a type 2 diabetes diagnosis code (ICPC-2 code T90 or ICD-10 code E11) The incident year of type 2 diabetes was defined as the earliest year in which these criteria were fulfilled, restricted to the study period (2009–2021). Prevalence was defined as any prior or concurrent incident year while still being alive and residing in Norway (also allowing onset before 2009).

### Covariates

Time trends in incidence and prevalence of type 2 diabetes during 2009–2021 were stratified by registered information on sex (male or female), date of birth (age), place of birth (continent, from Statistics Norway), and yearly maximum attained education level (<11 years, 11–13 years, and >13 years, from Statistics Norway).

### Data analysis

Data handling, incidence and prevalence estimation were done using R (v4.1.2) [10]. Incidence and prevalence estimates were presented both as unstandardised and direct standardised to the current study’s age distribution in one-year categories in 2021. We used Poisson regression to estimate the percent change in incidence rate over calendar years for the interrupted time series regression between the periods 2009–2014 and 2015–2021. We used JoinPoint (v4.9.1.0 with default settings except using the weighted Bayesian Information Criterion for model selection) to investigate possible trend breaks using a data driven method. P-values less than 5% were considered significant. We did four post hoc sensitivity analyses to better understand the time trends, as explained in the results section. See Electronic Supplemental Material (ESM) Methods for additional technical details and comments.

## Results

Across the study period 4 100 548 individuals aged 30–89 years were included, 80.2% were born in Norway, and 36.9% had more than 13 years of education in 2020 (Table 1).

**Table 1.** Characteristics of study population 30–89 years for analysis of incidence of type 2 diabetes from 1 January 2009 to 31 December 2021.

| Characteristic | N (%) | Incidence cases | Person years | Incidence rate per 100 000 |
| --- | --- | --- | --- | --- |
| All | 4,100,548<br>(100.0 %) | 195,935 | 38,478,517 | 509 |
| Male | 2,068,608<br>(50.4 %) | 114,875<br>(58.6 %) | 19,129,911<br>(49.7 %) | 600 |
| Female | 2,031,940<br>(49.6 %) | 81,060<br>(41.4 %) | 19,348,606<br>(50.3 %) | 418 |
| Age group* |  |  |  |  |
| 30–49 y | 2,433,029<br>(43.9 %) | 41,826<br>(21.3 %) | 17,741,294<br>(46.1 %) | 235 |
| 50–69 y | 2,016,629<br>(36.4 %) | 103,191<br>(52.6 %) | 14,553,649<br>(37.8 %) | 709 |
| 70–89 y | 1,084,398<br>(19.6 %) | 50,918 (26 %) | 6,183,574<br>(16.1 %) | 823 |
| Education level* |  |  |  |  |
| Low (<10 years) | 1,136,677<br>(27.1 %) | 67,970<br>(34.7 %) | 8,957,714<br>(23.3 %) | 758 |
| Middle (10–12 years) | 1,551,123<br>(36.9 %) | 84,372<br>(43.1 %) | 14,995,803<br>(39 %) | 562 |
| High (≥13 years) | 1,514,079 (36 %) | 43,593<br>(22.2 %) | 14,525,005<br>(37.7%) | 300 |
| Region of birth |  |  |  |  |
| Norway | 3,292,562<br>(80.3 %) | 160,793<br>(82 %) | 32,522,422<br>(84.5 %) | 494 |
| Europe | 464,858 (11.3 %) | 12,916<br>(6.6 %) | 3,442,949<br>(8.9 %) | 375 |
| Asia | 208,936 (5.1 %) | 14,870<br>(7.6 %) | 1,512,358<br>(3.9 %) | 983 |
| Africa | 78,925 (1.9 %) | 5,446 (2.8 %) | 573,846<br>(1.5 %) | 949 |
| North and Central America | 28,366 (0.7 %) | 912 (0.46 %) | 226,459<br>(0.6%) | 402 |
| South America | 23,297 (0.6 %) | 943 (0.48 %) | 176,433<br>(0.5 %) | 534 |
| Oceania | 3,601 (0.1 %) | 55 (0.03%) | 24,034<br>(0.06 %) | 228 |

\* = The total number of group entries exceeds the total number of unique participants because an individual's education level may change over the study period.

### Type 2 diabetes incidence trends

Throughout the study period, 195 935 individuals were registered with incident type 2 diabetes, and overall incidence rate declined from 651 in 2009 to 530 in 2021 per 100 000. After a decline in 2009–2013, the type 2 diabetes incidence rate stabilised (Figure 1). Interrupted time series regressions corroborated a change in the incidence decline during 2009–2014 (significant annual percent change = -7.3), which overall turned into a relatively small increase in 2015–2021 (significant annual percent change = 0.7). JoinPoint analysis showed trend-breaks in 2013 and 2019 (significant annual percent change 2009–2013 = - 7.9, 95% CI: -12.5, -3.0; annual percent change 2013–2019: -0.8, 95% CI: -4.7, 3.1; annual percent change 2019–2021: 7.0, 95%CI -9.5, 26.5). Males had higher type 2 diabetes incidence rate than females, but the incidence time trend patterns were similar in both sexes. Age-standardised time-trends were not markedly different from the unstandardised rates (Figure 1), but changes in the type 2 diabetes incidence rates were in absolute terms more pronounced in the older age groups (Figure 2). Age-groups 50–69 and 70–89 tended to converge from 2009–2015 with a greater absolute initial decline in the oldest age-group.

**Figure 1.**
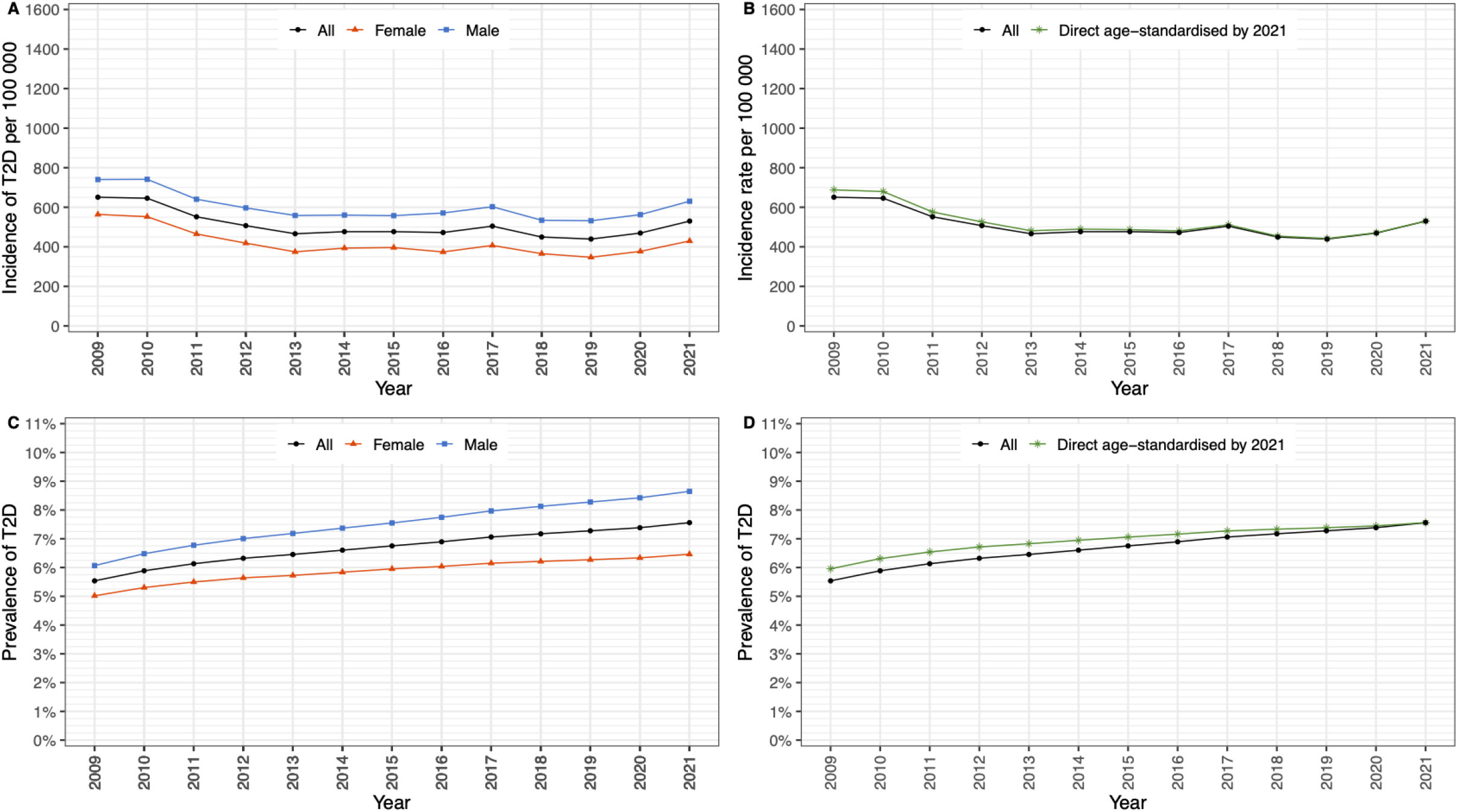
Type 2 diabetes incidence rate per 100 000 (top row; A & B) and type 2 diabetes prevalence (bottom row; C & D), grouped by left column: blue square = male, red triangle = female, black circle = all, and right column: without (black) and with direct age standardisation (green) using our current study population for the year 2021. Yearly age groups were used for standardisation.

**Figure 2.**
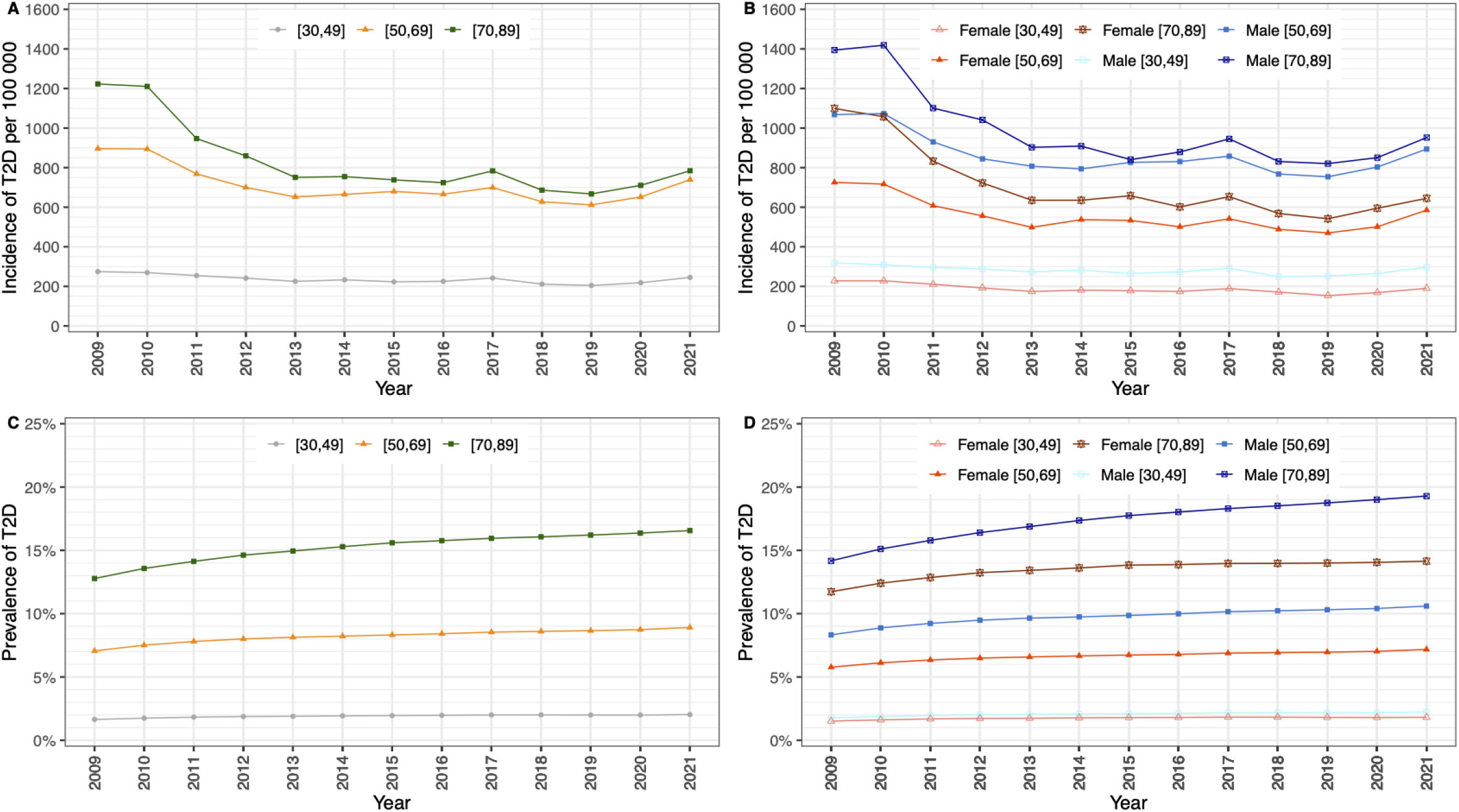
Type 2 diabetes incidence rate per 100 000 (top row; A & B) and type 2 diabetes prevalence (bottom row; C & D), grouped in left column: by only age group (grey circle = all [30,49>], orange triangle = all [50,69], dark green square = all [70,89]), and right column: by age group and sex (dark blue square= male [70,89], blue square = male [50,69], light blue square = male [30,49], dark red double triangle = female [70,89], red triangle = female [50,69], salmon triangle= female [30,49]).

Inhabitants born in Africa or Asia had higher incidence of type 2 diabetes, and inhabitants born in Europe outside Norway had a lower incidence than those born in Norway (Figure 3). Approximately twofold higher incidence rates were seen in individuals with lower education compared to those with higher education (Figure 3B). In absolute terms, the transient increase in 2017 and increase from 2019–2021 appeared to be more pronounced for individuals with lower education.

**Figure 3.**
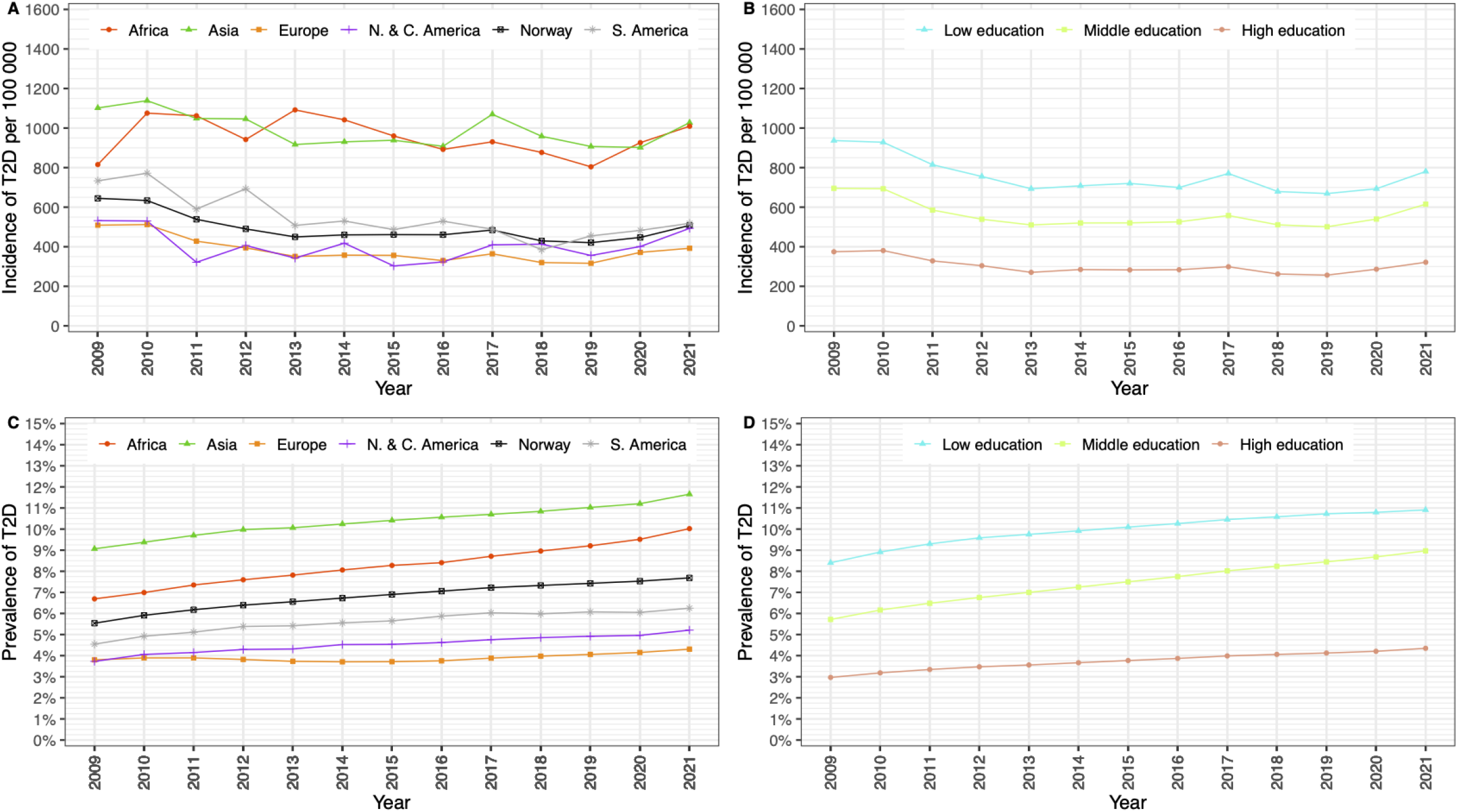
Type 2 diabetes incidence rate per 100 000 (top row; A & B) and prevalence (bottom row; C & D), grouped by left column: birth region (green triangle = Asia, red circle = Africa, black square = Norway, grey asterisk = South America, filled orange square = Europe (without Norway), purple cross = North and Central America), and by right column: years of education. Higher level of education corresponded to >13 years (salmon circle), middle level of education to 11-13 years (green square), and lower level of education to <11 years (light blue triangle). Oceania as birth region was not included due to too few observations.

### Type 2 diabetes prevalence

Overall, there were 165 432 prevalent cases (5.5% of the population) of type 2 diabetes in 2009, consistently increasing throughout the study period to a peak at 259 017 prevalent cases (7.5%) in 2021. Risk factors for prevalent type 2 diabetes were the same as that for incident type 2 diabetes (Figures 1-3). The prevalence accelerated more in males (Fig. 1C) and in the group with medium level education (Figure 3D). The prevalence for most study years peaked around age 80 and tended to plateau towards the end of the study at 17-18% after age 75 (ESM Fig 1).

### Sensitivity analyses

First, we investigated trends separately for those who had used non-insulin glucose-lowering drugs at least once and for those that had not. We found very similar trajectories over time, except that the transient increase in incidence in 2017 was absent in those who did not use non-insulin glucose-lowering drugs (ESM Fig. 2). Second, excluding SGLT-2 inhibitors from the definition of type 2 diabetes gave near identical results (ESM Fig. 3). Third, excluding GLP-1 agonists from the definition of type 2 diabetes also resulted in near identical curves (ESM Fig 4). Fourth, we restricted the time-window between the first two registrations needed to fulfil the type 2 diabetes definition to a maximum of two years to investigate whether a changing time-window could explain some of the time trends. While the absolute estimates were lower with this restriction, as expected, incidence and prevalence trends over time appeared similar (ESM Fig. 5). Finally, we plotted trends in annual mortality to investigate whether the increasing prevalence, despite a relatively stable incidence rate, could be explained by decreasing mortality rates among prevalent cases compared to the general population. Annual mortality among those with type 2 diabetes declined slightly over time, in parallel with the mortality among those without type 2 diabetes (ESM Fig. 6).

## Discussion

The decline in type 2 diabetes incidence from 2009–2014 was not sustained, while the prevalence continued to increase throughout the period 2009–2021. Absolute incidence rates and prevalences were higher for those with lower education level and for those born in Africa or Asia, but overall time trends were largely consistent between groups.

### Comparison of time trends in other studies

Incidence of type 2 diabetes declined or stabilised in many high-income countries from around 2010 onwards [1, 2]. A Danish study with data for 1995–2018 showed increasing incidence rate until 2011 followed by a decline [11]. In Bavaria, Germany, declining incidence was observed during 2012–2021, with strong decreases only until 2017 [12]. In France among the pharmacologically treated, a nationwide decline in incidence was observed in 2013–2019, followed by an increasing incidence in 2020–2021 [13]. Scottish surveys similarly showed an increase in 2021–2023 in type 2 diabetes incidence rates [14, 15], while declining rate was reported in the period 2010–2019 from Australia [9].

In our study, we observed stable incidence and prevalence trends in the age-group 30–49 years, and the Bavarian study showed similar stable crude incidence rates for the same age [12]. The 30–49-year-olds in the Danish study showed an increase in standardised incidence rates until 2011, followed by a slight decrease [11]. Australian data showed decreasing incidence for all age-groups [9]. An increasing incidence during 2019–2021 was also observed in the Bavarian and the French studies [12, 13].

Low socio-economic status was consistently associated with higher incidence of type 2 diabetes in our and in other studies [8, 9]. Time trends were largely parallel in groups of different area-based socioeconomic status in studies from Scotland and Australia, except a tendency of a widening inequality in Scotland 2004–2013 [8, 9]. In our study, the prevalence among inhabitants with middle level education tended to increase somewhat more than for the two other educational levels. Inhabitants in our study born in Asia and Africa had higher absolute incidence rates of type 2 diabetes compared to the other visualised birth regions, but time trends were still largely consistent across birth region groups. In contrast, in Australia, immigrants born in Asia, North Africa and the Middle East had increasing incidence trends while the other groups had stable or declining trends during 2013–2019 [9].

### Possible explanations for the time trends

To explain the common declining type 2 diabetes incidence trends around 2009–2014, several explanations have been suggested: the possibility of a true decline due to preventive interventions and due to changing diagnostic criteria and activity over time [2, 16]. The stabilising incidence rate in Norway after 2013 may support the hypothesis that the previous decline was due to changing diagnostic criteria and intensified case-finding, although firm conclusions cannot be drawn. We lack nationwide data on undiagnosed diabetes in Norway, but The HUNT survey showed that undiagnosed cases (defined by HbA1c at 6.5% or more (48 mmol/mol) without a previous diagnosis) comprised 11% of all diabetes cases in the Nord-Trøndelag region in 2017-2019 [17]. In The Tromsø study in the municipality of Tromsø, the proportion of all diabetes cases that were undiagnosed according to HbA1c declined from 41.8%–51.4% in 2001, via 31.8%–35.2% in 2006–2007, to 17.2%–24.2% in 2015–2016 [18]. This decrease over 15 years in proportion of undiagnosed cases also support the notion that our decline in incidence may be due to intensified case-finding. However, we neither know incidence before 2009 nor cannot exclude the possibility that the repeated waves of health screening in Tromsø municipality may itself have partially depleted the population of undiagnosed cases of diabetes that would not have been seen in the rest of the Norwegian population with no such screening. Without nationally representative and long-term data on diagnostic activity it is hard to draw firm conclusions. Body mass index and central adiposity are strong determinants of type 2 diabetes, and mean body mass index, waist circumference and body fat increased over time in adult participants in the Tromsø study until the most recent survey in 2016 [19-21]. This seems to contrast with a true declining or flat incidence curve.

Why has the prevalence increased while the incidence decreased and then largely stabilised? Theoretically, increases in prevalence may be explained by population ageing resulting from improved diabetes treatment and subsequent increased longevity. Our mortality data for those with type 2 diabetes support the notion of increased longevity, with overall decreasing mortality parallel to the general study population. Furthermore, age-standardisation did not affect incidence rates notably. We therefore believe that the increasing prevalence is largely due to increasing incidence before our current study period, combined with improved longevity. Prevalence trends are slower to change than incidence, because the long average duration of diagnosed type 2 diabetes and because the number of incident cases during a year is small compared to the number of prevalent cases.

Finally, we can speculate about the tendency towards an increasing incidence from 2019– 2021. This may be a true effect, partially explained by multiple changes in behaviour during the corona pandemic from 2020 [22]. Increasing use of incretin mimetics such as GLP-1 receptor agonists for weight loss [23], and possible off-label use can in theory also influence the apparent increase, although our sensitivity analysis suggested otherwise.

### Strengths and limitations

The main strength of our study is our use of nationwide register-linkage with a long-time follow-up period. The classification relies on diagnostic codes used by clinicians in primary health care and hospitals (including inpatient and outpatient care), and codes on drug dispensation from the pharmacies, together reducing the risk of misclassification. Our use of individual level data on educational level and country of birth is a strength, while the lack of information about undiagnosed diabetes and adiposity are limitations.

## Conclusions

In conclusion, the previously reported decline in incidence of diagnosed type 2 diabetes from 2009–2014 was not sustained, as incidence of type diabetes was relatively stable from 2015–2021, with a suggestive increase during 2019–2021. The prevalence continued to increase.

## Data Availability

Data are accessible to authorised researchers after ethical approval and application to https://helsedata.no/en/, administered by the Norwegian Institute of Public Health.

## Author contributions

Conceptualisation: HLG, EQ, LCS, GT, PLDR; Methodology: LCS, GT, PLDR, IJB, LBH; Writing-Original Draft: LCS, PLDR, LBH; Review & Editing: HLG, EQ, LCS, GT, PLDR, IJB, LBH; Visualisation: LBH.

## Acknowledgments

The authors would like to thank Øystein Karlstad and Vidar Hjellvik for help with initial data curation.

## References

[1] Magliano DJ, Chen L, Islam RM, et al. (2021) Trends in the incidence of diagnosed diabetes: a multicountry analysis of aggregate data from 22 million diagnoses in high-income and middle-income settings. Lancet Diabetes Endocrinol 9: 203–211

[2] Magliano DJ, Islam RM, Barr ELM, et al. (2019) Trends in incidence of total or type 2 diabetes: systematic review. BMJ 366: l5003

[3] Ruiz PLD, Stene LC, Bakken IJ, Håberg SE, Birkeland KI, Gulseth HL (2018) Decreasing incidence of pharmacologically and non-pharmacologically treated type 2 diabetes in Norway: a nationwide study. Diabetologia 61: 2310–2318

[4] Kalyani RR, Neumiller JJ, Maruthur NM, Wexler DJ (2025) Diagnosis and treatment of type 2 diabetes in adults: a review. JAMA 334: 984–1002

[5] Ahmad E, Lim S, Lamptey R, Webb DR, Davies MJ (2022) Type 2 diabetes. Lancet 400: 1803–1820

[6] NCD Risk Factor Collaboration (2016) Trends in adult body-mass index in 200 countries from 1975 to 2014: a pooled analysis of 1698 population-based measurement studies with 19.2 million participants. Lancet 387: 1377–1396

[7] GBD Adult BMI Collaborators (2025) Global, regional, and national prevalence of adult overweight and obesity, 1990-2021, with forecasts to 2050: a forecasting study for the Global Burden of Disease Study 2021. Lancet 405: 813–838

[8] Read SH, Kerssens JJ, McAllister DA, et al. (2016) Trends in type 2 diabetes incidence and mortality in Scotland between 2004 and 2013. Diabetologia 59: 2106–2113

[9] Magliano DJ, Chen L, Morton JI, Buyadaa O, Salim A, Shaw JE (2024) Changes in the incidence of type 2 diabetes in Australia, 2005-2019, overall and by socio-demographic characteristics: a population-based study. Med J Aust 221: 473–479

[10] R Team (2021) A language and environment for statistical computing

[11] Knudsen JS, Knudsen SS, Hulman A, et al. (2022) Changes in type 2 diabetes incidence and mortality associated with introduction of HbA1c as diagnostic option: A Danish 24-year population-based study. Lancet Reg Health Eur 14: 100291

[12] Lehner CT, Eberl M, Donnachie E, et al. (2024) Incidence trend of type 2 diabetes from 2012 to 2021 in Germany: an analysis of health claims data of 11 million statutorily insured people. Diabetologia 67: 1040–1050

[13] Fosse-Edorh S, Guion M, Goria S, Mandereau-Bruno L, Cosson E (2025) Dynamics of diabetes prevalence, incidence and mortality in France: A nationwide study, 2013-2021. Diabetes Metab 51: 101615

[14] Scottish Diabetes Group (2024) Scottish Diabetes Survey 2023. Available from https://www.diabetesinscotland.org.uk/wp-content/uploads/2024/11/Scottish-Diabetes-Survey-2023.pdf, accessed June 2025

[15] Scottish Diabetes Group (2021) Scottish Diabetes Survey 2020. Available from https://www.diabetesinscotland.org.uk/wp-content/uploads/2022/01/Diabetes-Scottish-Diabetes-Survey-2020.pdf, accessed June 2025

[16] Selvin E, Ali MK (2017) Declines in the incidence of diabetes in the U.S.-real progress or artifact? Diabetes Care 40: 1139–1143

[17] Bjarkø VV, Haug EB, Sørgjerd EP, et al. (2022) Undiagnosed diabetes: Prevalence and cardiovascular risk profile in a population-based study of 52,856 individuals. The HUNT Study, Norway. Diabet Med 39: e14829

[18] Langholz PL, Wilsgaard T, Njølstad I, Jorde R, Hopstock LA (2021) Trends in known and undiagnosed diabetes, HbA1c levels, cardiometabolic risk factors and diabetes treatment target achievement in repeated cross-sectional surveys: the population-based Tromso Study 1994–2016. BMJ Open 11: e041846

[19] Jacobsen BK, Aars NA (2015) Changes in body mass index and the prevalence of obesity during 1994–2008: repeated cross-sectional surveys and longitudinal analyses. The Tromsø Study. BMJ Open 5: e007859

[20] Løvsletten O, Jacobsen BK, Grimsgaard S, et al. (2020) Prevalence of general and abdominal obesity in 2015–2016 and 8-year longitudinal weight and waist circumference changes in adults and elderly: the Tromsø Study. BMJ Open 10: e038465

[21] Lundblad MW, Johansson J, Jacobsen BK, et al. (2021) Secular and longitudinal trends in body composition: The Tromsø Study, 2001 to 2016. Obesity 29: 1939–1949

[22] Zhang T, Mei Q, Zhang Z, et al. (2022) Risk for newly diagnosed diabetes after COVID-19: a systematic review and meta-analysis. BMC Med 20: 444

[23] Ruiz PL, Hindenes LB, Karlstad O, et al. (2025) Trends in the use of drugs with weight-loss effect: Scandinavian study from 2017 to 2023. Diabetes Obes Metab 27: 2901–2905

