## Supplemental material for "Type 2 diabetes in Norway 2009–2021: Have declining incidence rates continued?"

#### Electronic Supplementary Material (ESM)

### ESM Methods

*Additional details of data and analysis*

Figures were created using the “ggplot2” package and the “gridExtra” package. We used the “data.table” package to read and write large data files. Program code can be requested from the authors. Due to the data resolution being yearly, i.e. we have no measurement of exact entry- or exit days, individuals could in theory enter the study the last few days of a year, or exit the first few days of a year, and still be considered included that study year. We defined being 30 years old as for example having a registered birth year as 1991 and then being 30 years old in 2021; i.e. current study year minus the birth year. Only a single entry by immigration was allowed per individual. A valid registry observation was defined as the “regstatus” variable being equal to “1” (resident) for a respective study year. The following URL describes this “regstatus” variable from SSB in detail: https://www.ssb.no/a/metadata/codelist/datadok/1276589/en.

Incidence and prevalence estimates were direct standardized to the current study’s yearly age distribution in 2021. Original Norwegian education codes were either missing, IU (0 years, unknown or not completed any education), GS (1-10 first years, equivalent to primary school, middle or secondary school, or just up until high school), VGS (11-13 years, similar to high school and upper secondary school), UHK and FS (>13 years, any additional education, including bachelor's degree or shorter) and UHL (>13 years, further higher education, including master's degree or longer). Inhabitants (N = 166 146, 4%) with missing information about maximum attained education were imputed as having less than 11 years of education for consistency with the IU code. We did not have information about education level after 2020. URL to documentation: https://www.ssb.no/klass/klassifikasjoner/225/koder. Missing information about place of birth was not imputed, and the three inhabitants affected were instead excluded from any analysis including such grouping.

### ESM Figures

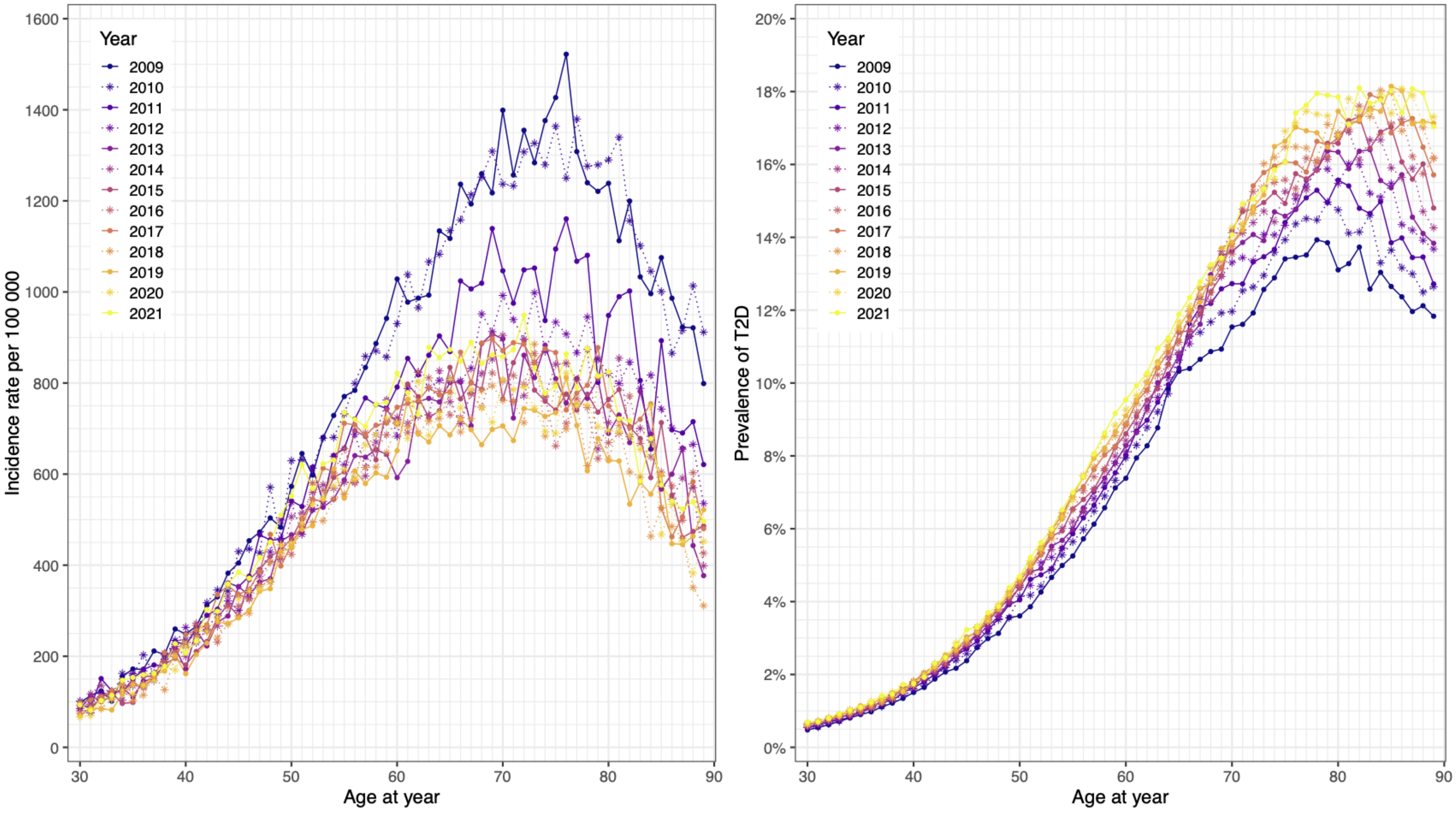

ESM Fig. 1: Type 2 diabetes (A) incidence and (B) prevalence of type 2 diabetes by age, stratified by calendar year.

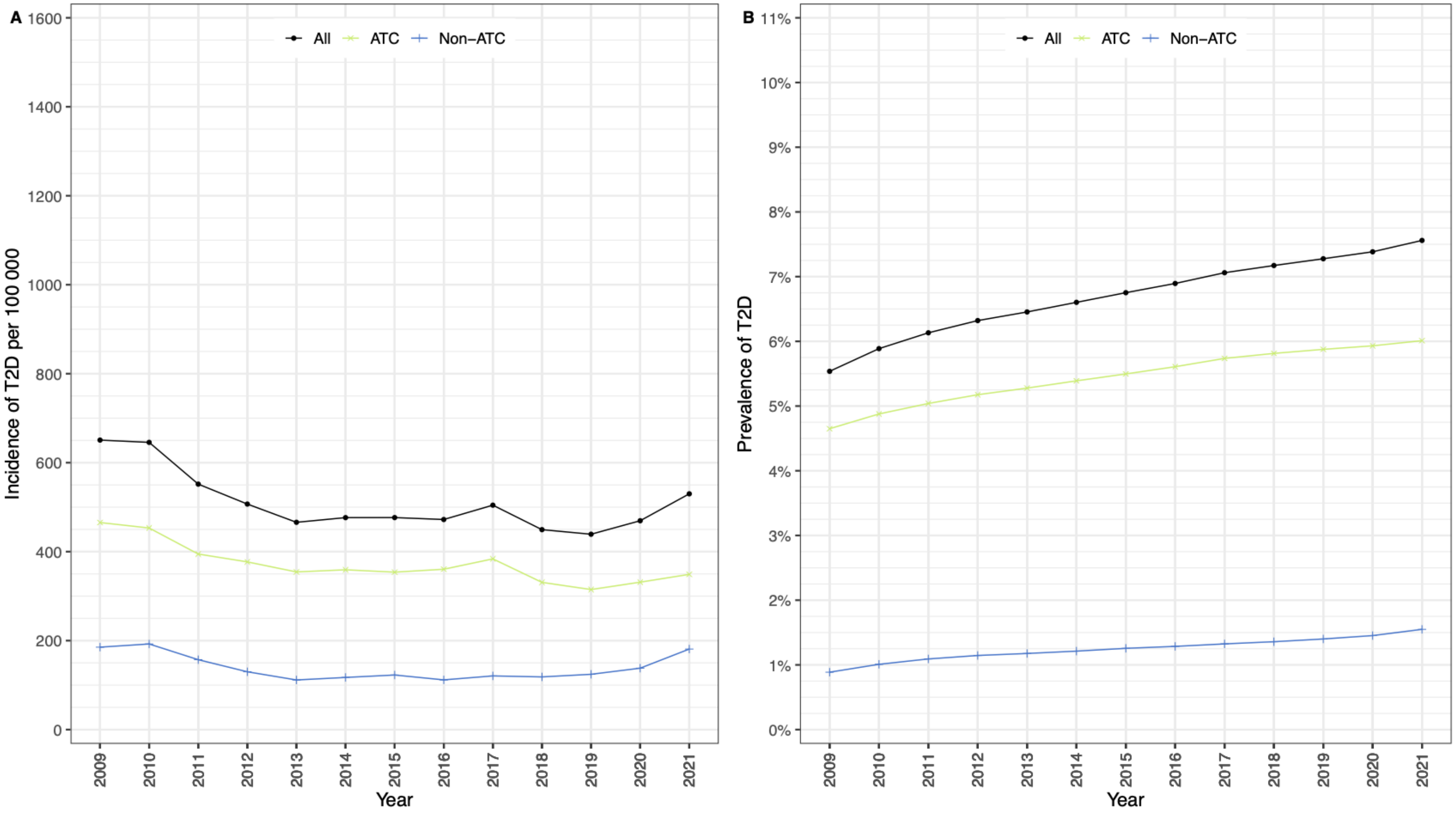

ESM Fig. 2. Sensitivity analysis: Trends in (A) incidence and (B) prevalence of type 2 diabetes in people with (ATC) or without (non-ATC) at least one prescribed non-insulin glucose lowering drug (ATC: Anatomic Therapeutic Chemical, only codes starting with A10B for non-insulin glucose lowering drugs).

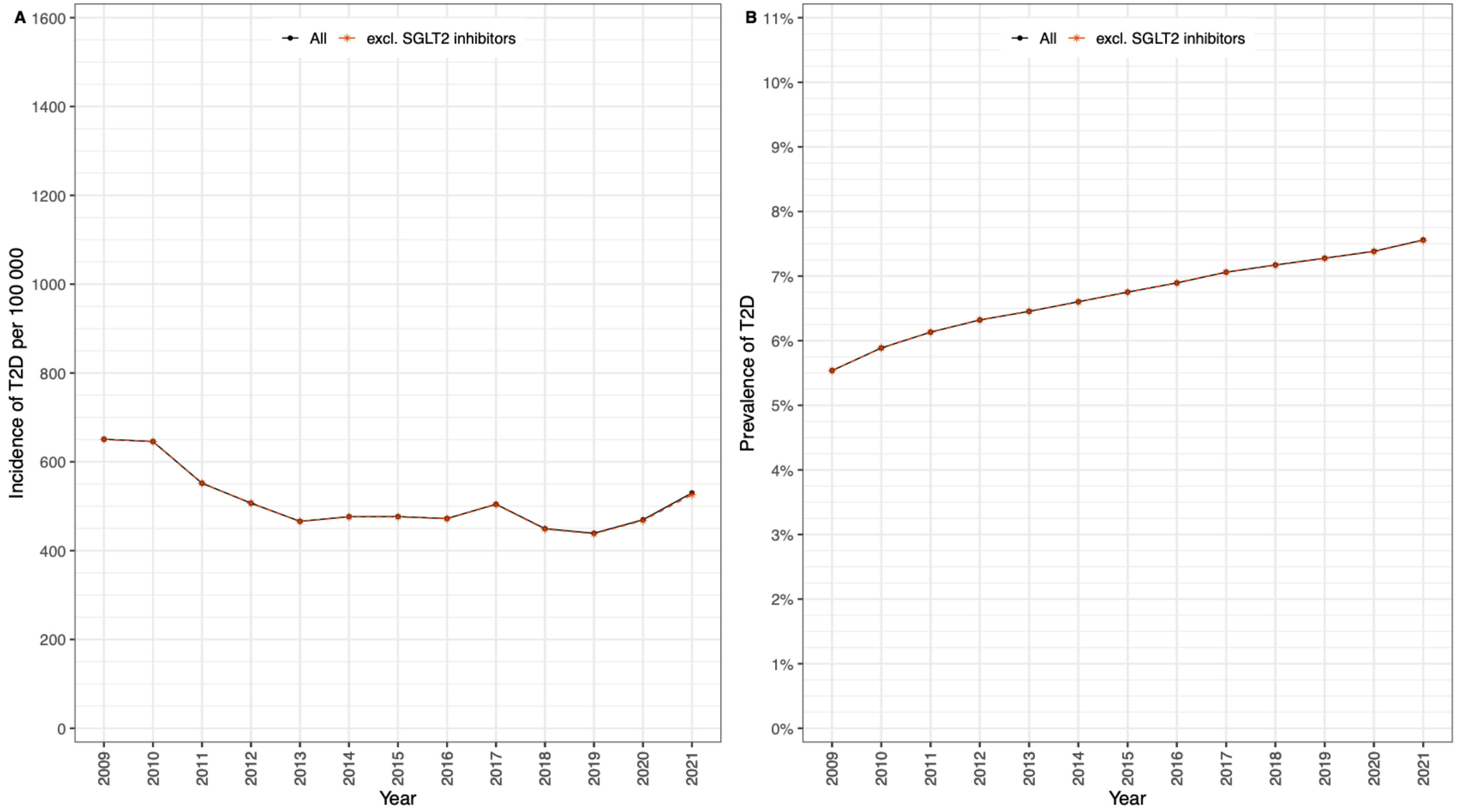

ESM Fig. 3. Sensitivity analysis: Trends in (A) incidence and (B) prevalence of type 2 diabetes after excluding SGLT2 inhibitors from the diagnostic algorithm. Note that the two curves (all and SGLT2 inhibitors excluded) are nearly completely superimposed, illustrating that this had no notable effect on observed trends. Sodium-Glucose co-transporter 2 (SGLT2) inhibitors, Gliflozins, ATC code A10BK).

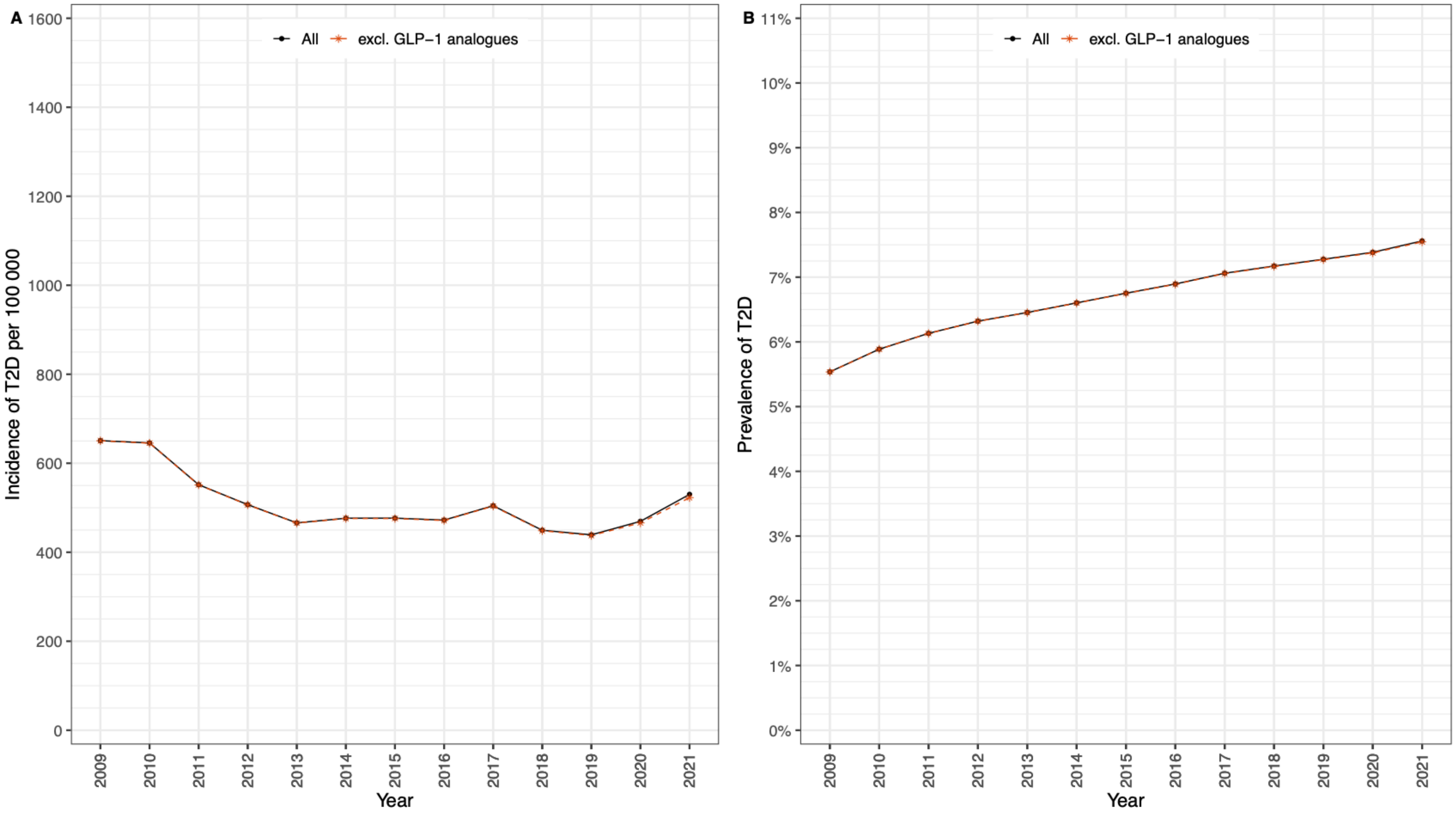

ESM Fig. 4. Sensitivity analysis: Trends in type 2 diabetes (A) incidence and (B) prevalence after excluding GLP-1 analogues (ATC code A10BJ) from the diagnostic algorithm. Note also here that the two curves (all and GLP-1 agonists excluded) are nearly superimposed except for after 2019.

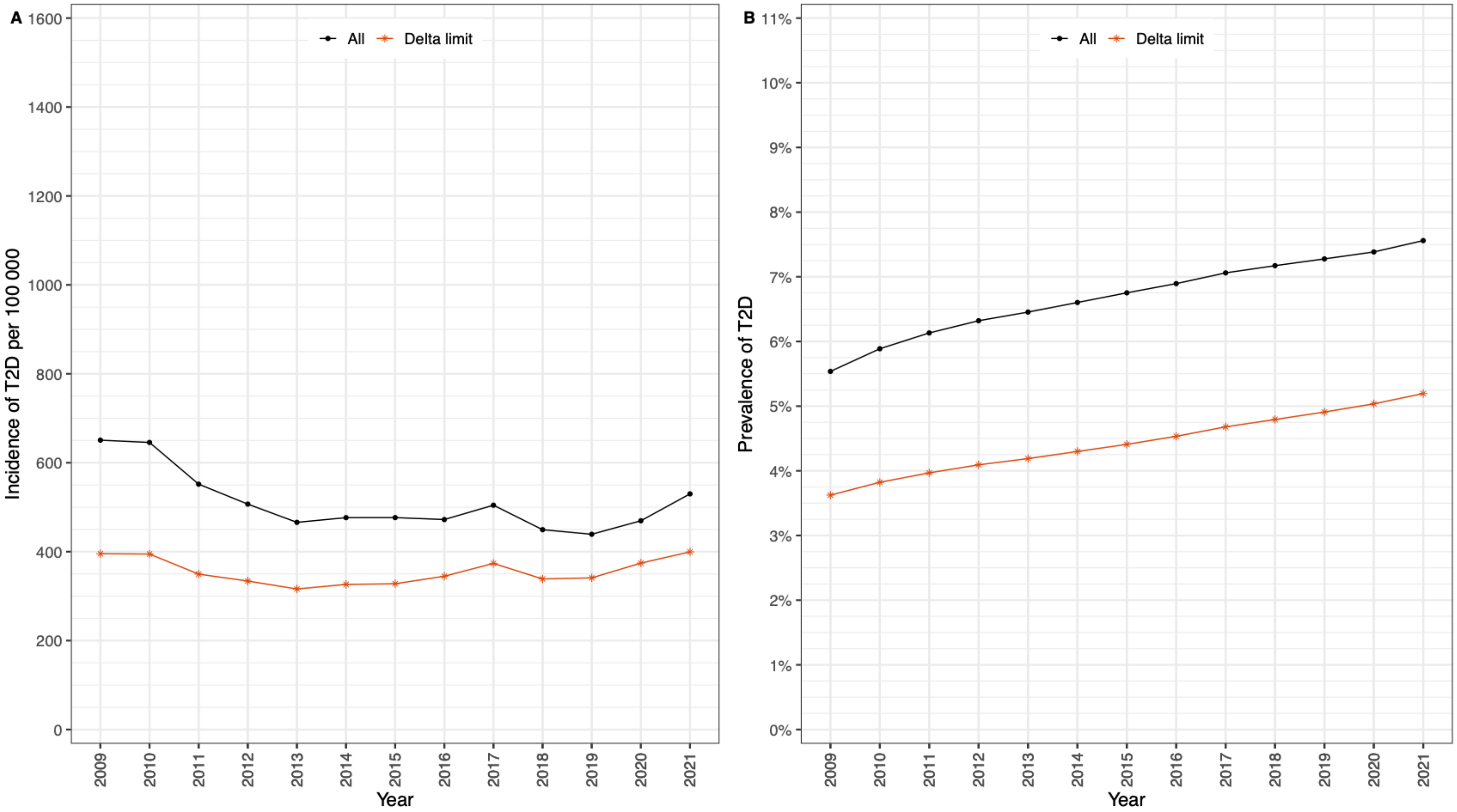

ESM Fig. 5. Sensitivity analysis: Type 2 diabetes (A) incidence and (B) prevalence trends after limiting the time between the first and second registry observation (i.e. delta limit) to a maximum of two years.

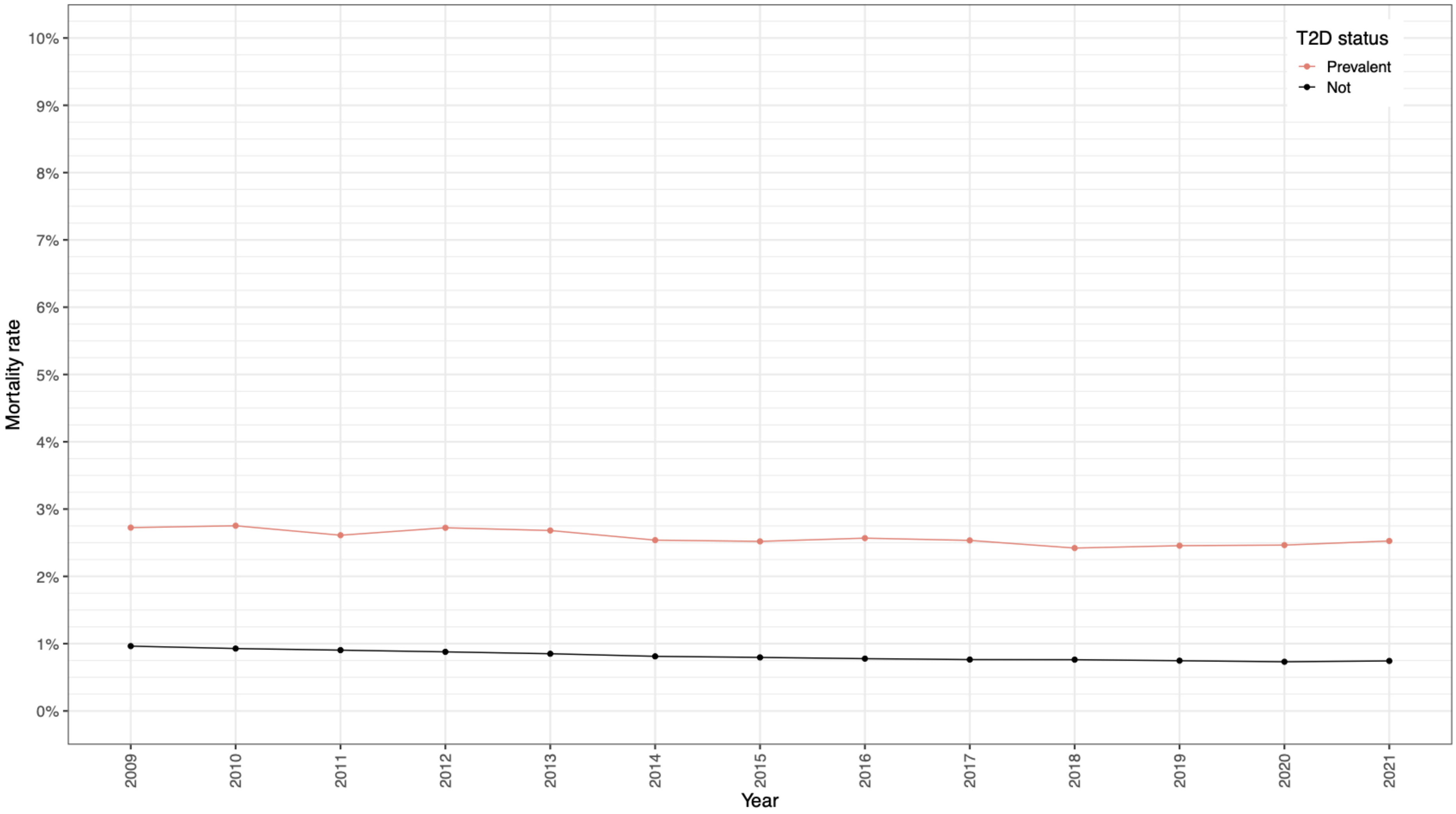

ESM Fig. 6. Annual mortality in people aged 30-89 years with and without prevalent type 2 diabetes, by calendar year.
